# Risk of Hospitalization, severe disease, and mortality due to COVID-19 and PIMS-TS in children with SARS-CoV-2 infection in Germany

**DOI:** 10.1101/2021.11.30.21267048

**Authors:** AL Sorg, M Hufnagel, M Doenhardt, N Diffloth, H Schroten, R v. Kries, R Berner, J Armann

## Abstract

**Background:** Although children and adolescents have a lower burden of SARS-CoV-2-associated disease as compared to adults, assessing absolute risk among children remains difficult due to a high rate of undetected cases. However, without more accurate case numbers, reliable risk analyses are impossible.

**Methods:** We combine data from three sources — a national seroprevalence study (the SARS-CoV-2 KIDS study), the German statutory notification system and a nationwide registry on children and adolescents hospitalized with either SARS-CoV-2 or Pediatric Inflammatory Multisystem Syndrome (PIMS-TS) — in order to provide reliable estimates on children’s hospitalization, intensive care admission and death due to COVID-19 and PIMS-TS.

**Results:** While the overall hospitalization rate associated with SARS-CoV-2 infection was 35.9 per 10,000 children, ICU admission rate was 1.7 per 10,000 and case fatality was 0.09 per 10,000. Children without comorbidities were found to be significantly less likely to suffer from a severe or fatal disease course. The lowest risk was observed in children aged 5-11 without comorbidities. In this group, the ICU admission rate was 0.2 per 10,000 and case fatality could not be calculated, due to an absence of cases. The overall PIMS-TS rate was 1 per 4,000 SARS-CoV-2 infections, the majority being children without comorbidities.

**Conclusion:** Overall, the SARS-CoV-2-associated burden of a severe disease course or death in children and adolescents is low. This seems particularly the case for 5-11-year-old children without comorbidities. By contrast, PIMS-TS plays a major role in overall disease burden among all pediatric age groups.

## Introduction

Since December 2019, Severe Acute Respiratory Syndrome Coronavirus 2 (SARS-CoV-2) [1] has rapidly spread worldwide; by March 2020 the World Health Organization (WHO) had declared it a pandemic. As compared to adults [2], the overwhelming evidence demonstrates that children and adolescents usually have mild disease courses, along with low disease-associated morbidity and mortality [3–5]. Nevertheless, their absolute risk remains difficult to assess. Risk assessment requires accurate case numbers and valid estimates of the population at risk. Due to a high rate of undetected cases among children [6], cases reported to the statutory notification systems significantly underestimate overall infections in this age group. Therefore, unless corrected accordingly, this can lead to a substantial overestimation of the overall risk for children and adolescents. Additionally, a relevant number of patients with laboratory-confirmed SARS-CoV-2 infections during a hospital stay for different reasons may inflate the number of cases and thus the risk assessment even further [7].

Without reliable data on actual infection-associated disease burden, a meaningful risk-benefit assessment of pandemic control and mitigation measures affecting this particular age group is impossible.

Here, we analyze information from a seroprevalence study in children – the SARS-CoV-2 KIDS study, recently published by Sorg et al. [8] – with publicly accessible data from the German statutory notification system on hospitalization and mortality [9], and data from a nationwide registry on hospitalized children with COVID-19 and Pediatric Inflammatory Multisystem Syndrome - Temporarily-Associated with SARS-CoV-2 (PIMS-TS), the latter of which is sponsored by the German Society for Pediatric Infectious Diseases (DGPI) [10,11]. By combining this data, we have aimed to provide a validated risk assessment regarding hospitalization, intensive care admission and death due to COVID-19 and PIMS-TS for children of different age groups. The choice of age groups was made based upon vaccination eligibility in Germany.

## Methods

Our analysis is based upon information extracted from the results of three different surveys.

### SARS-CoV-2 KIDS study

The proportion of children with positive titers against SARS-CoV-2 immunoglobulin G (IgG) is based upon recently published results from the SARS-CoV-2 KIDS study [8]. This was a hospital-based, multi-center, longitudinal study in children aged ≤17 years. From June 2020 to May 2021, in 14 pediatric hospitals across Germany, 10,358 participants were recruited during their inpatient or outpatient stays, (monthly average recruiting rate: 863 (±SD 220) participants). Blood samples taken for routine clinical procedures were additionally tested for the IgG-specific S1 domain of SARS-CoV-2 spike protein by means of an Enzyme-Linked Immunosorbent Assay (ELISA - Euroimmun Medizinische Diagnostika AG, Lübeck, Germany). As generally recommended by the manufacturer, an optical density ratio above 1.1 defined sera samples as anti-SARS-CoV-2 IgG positive.

The SARS-CoV-2 KIDS study authors reported a seroprevalence of SARS-CoV-2 IgG antibodies of 10.8% (95% CI 8.7, 12.9) for March 2021, and observed no major change through May 2021 [8]. Additionally, by the end of the observation period, they detected no differences in seroprevalence among the different age groups.

The German Federal Statistical Office provides updated data on the total population of children in Germany and the breakdown of age groups [12]. The seroprevalence estimates included in the SARS-CoV-2 KIDS study publication were extrapolated for the total population.

### DGPI Registries

In March 2020, near the beginning of the pandemic, a national, prospective registry for children and adolescents hospitalized with a SARS-CoV-2 infection in Germany was established. On May 28, 2020, it was expanded to also capture PIMS-TS cases. All German pediatric hospitals, along with members of the German scientific pediatric and pediatric infectious diseases societies, (German Society of Pediatric Infectious Diseases (DGPI), German Society of Pediatrics and Adolescents Medicine (DGKJ), Association of Senior Pediatricians and Pediatric Surgeons in Germany (VLKKD)), were invited to participate. Included for analysis were hospitalized patients under 17 years old who had laboratory-confirmed SARS-CoV-2 infections, (reported to the registry from March 1, 2020 - May 31, 2021), as well as those who fulfilled the WHO case definition for PIMS-TS [13], (reported to the registry from May 28, 2020 - May 31, 2021).

Via a link on the DGPI website (https://dgpi.de/covid-19-survey-der-dgpi/), access was provided to an electronic case report form on a REDCap (Research Electronic Data Capture) platform hosted at Technische Universität Dresden. REDCap is a secure, web-based software platform designed to support data capture for research studies, providing 1) an intuitive interface for validated data capture; 2) audit trails for tracking data manipulation and export procedures; 3) automated export procedures for seamless data downloads to common statistical packages; and 4) procedures for data integration and interoperability with external sources. Data collected included demographic characteristics, symptoms and clinical signs, treatments, disease course during hospitalization and outcome at hospital discharge.

### Statutory notification system

The third survey included publicly available data from the statutory notification system of laboratory-confirmed SARS-CoV-2 infections in Germany [9]. In Germany, laboratory confirmation requires detection of SARS-CoV-2 nucleic acid by PCR or by culture isolation of the pathogen (according to the national case definition [14]). Laboratories are required to report cases to the local public health authorities (PHA), who then forward the information via the respective state PHA to the Robert Koch-Institute (RKI), (Germany’s national public health institute), in Berlin. Reporting includes information on hospitalization status and mortality. Relevant data are made publicly available via the website of the Robert Koch-Institute (www.rki.de). In our analysis, we referenced data of patients 0-14 years of age, reported between March 18, 2020 and May 31, 2021. To estimate the coverage extent of the DGPI registry, we compared the numbers reported from the statutory notification system with those from the DGPI registry regarding hospitalized children aged 0-14 years.

### Outcomes of interest

To assess the morbidity and mortality risk to children infected with SARS-CoV-2, we primarily were interested in the following six outcome measurements:

- Hospitalization associated with laboratory-confirmed SARS-CoV-2 infection: The reason for hospitalization was not always the patient’s SARS-CoV-2 infection. In some cases, the infection was detected during an inpatient stay for another medical reason. We did not exclude cases if or when additional concurrent infections other than SARS-CoV-2 (such as viral or bacterial pneumonia or bacterial non-pulmonary infections) were reported. This was the case in 10% of all reported cases. To compensate for underreporting, we adjusted the number of hospitalizations based upon the information regarding laboratory-confirmed SARS-CoV-2 infections in the statutory notification system.
- COVID-19 requiring therapy: The number of hospitalized patients requiring any form of therapeutic intervention for COVID-19, (as defined by the physician who reported the case), also was adjusted for underreporting.
- COVID-19 requiring ICU admission: The number of patients admitted to intensive care units (ICUs) due to COVID-19 was adjusted for possible underreporting.
- Death associated with COVID-19: The number of case fatalities due to COVID-19 was based upon the total reported in the statutory notification system.
- Hospitalization associated with PIMS-TS: Patients hospitalized due to PIMS-TS reported in DGPI registry.
- ICU stay associated with PIMS-TS: Patients admitted to intensive care unit (ICU) due to PIMS-TS
- Death associated with PIMS-TS: Case fatality due to PIMS-TS.

### Statistical Analysis

To calculate the risk for the different outcome measurements, we estimated the number of SARS-CoV-2-infected children by using the number of children in the respective age group as reported by the Federal Statistical Office, along with the proportion of IgG-seropositive children as reported in the SARS-CoV-2 KIDS Study by Sorg et al. [8]. To account for the uncertainty of this proportion, the upper and lower limits of the reported 95% confidence intervals served as a denominator for the risk estimate limits regarding the SARS-CoV-2 outcome measurements.

In contrast to the DGPI registry reporting, which is voluntary, reporting to the statutory notification system is compulsory under German law. To account for underreporting to the DGPI registry, we multiplied the numbers for hospitalization, COVID-19 patients requiring therapeutic interventions and ICU admissions due to COVID-19 by the rate of underreporting. Underreporting in turn was calculated by dividing the hospital cases published by the statutory reporting system with the numbers reported to the DGPI registry. The statutory notification system only provides data for the age group 0-14. From March 2020 to May 2021, 1,226 children aged 0-14 years old were reported in the DGPI registry. During the same period, the statutory notification system recorded 4,035 cases. This allowed us to calculate an underreporting rate of 3.29 (4,035/1,226).

We did not adjust the overall number of PIMS-TS cases, because these cases are not reported in the statutory health system. To date, there has been no other source for data on PIMS-TS in Germany. For this reason, it is not currently possible to evaluate a more precise coverage rate for the PIMS-TS registry.

The number of children living in Germany without comorbidities is unknown. Some information on medical history was available for the cases reported via the DGPI registry. To estimate the outcome risks for the children for whom we had no medical history, we reduced the population size by the proportion of children with comorbidities, as reported in the DGPI registries. Information on comorbidities was available for 1,602 children and was reported for 462 (28.8%) of them. On this basis, we postulated that 28.8% of the relevant population would have relevant comorbidities, most likely overestimating the proportion in the general pediatric population, because infected children with risk factors are more likely to be hospitalized. The age range groupings were selected in response to current discussions regarding COVID-19 vaccination recommendations for children in Germany. In Germany, vaccination against SARS-CoV-2 is recommended for the 12-17 age group. At the time of this writing, a vaccine has been licensed for 5–11-year-old children, but no official recommendation has been made. For younger children, a vaccine has yet to be licensed.

Chi-Square tests were used to examine the differences between the age groups and between the groups with and without comorbidities.

Our analysis was performed using SAS Version 9.4 (SAS Institute, Cary, NC, USA).

### Ethics

The DGPI registry was approved by the Ethics Committee of the Technische Universität (TU) Dresden (BO-EK-110032020) and was assigned clinical trial number DRKS00021506.

All other (anonymized) data was publicly available for scientific purposes.

## Results

As of December 31, 2020, approximately 13.7 million people in Germany were <18 years old [12]. With a seroprevalence of 10.8% (95 CI 8.7, 12.9) in May 2021, (as estimated by the SARS-CoV-2 KIDS study), and without substantial differences according to age group, an analysis of the combined data shows that 1,484,346 (1,195,723; 1,772,969) children likely already have been in contact with the virus. As such, they form a population at-risk for hospitalization, therapeutic interventions, ICU admission and/or death due to COVID-19 and/or PIMS-TS.

The flowchart displays the reported number of the respective outcomes (n) and the numbers adjusted for underreporting (n_c_), where appropriate. In the 5-11-year-old age group, 89 cases required SARS-CoV-2-related therapy. In the 12-17-year-old group, the number of cases was 352. In both age groups, the absolute numbers were markedly reduced when limited to cases requiring intensive care unit admission (Figure 1).

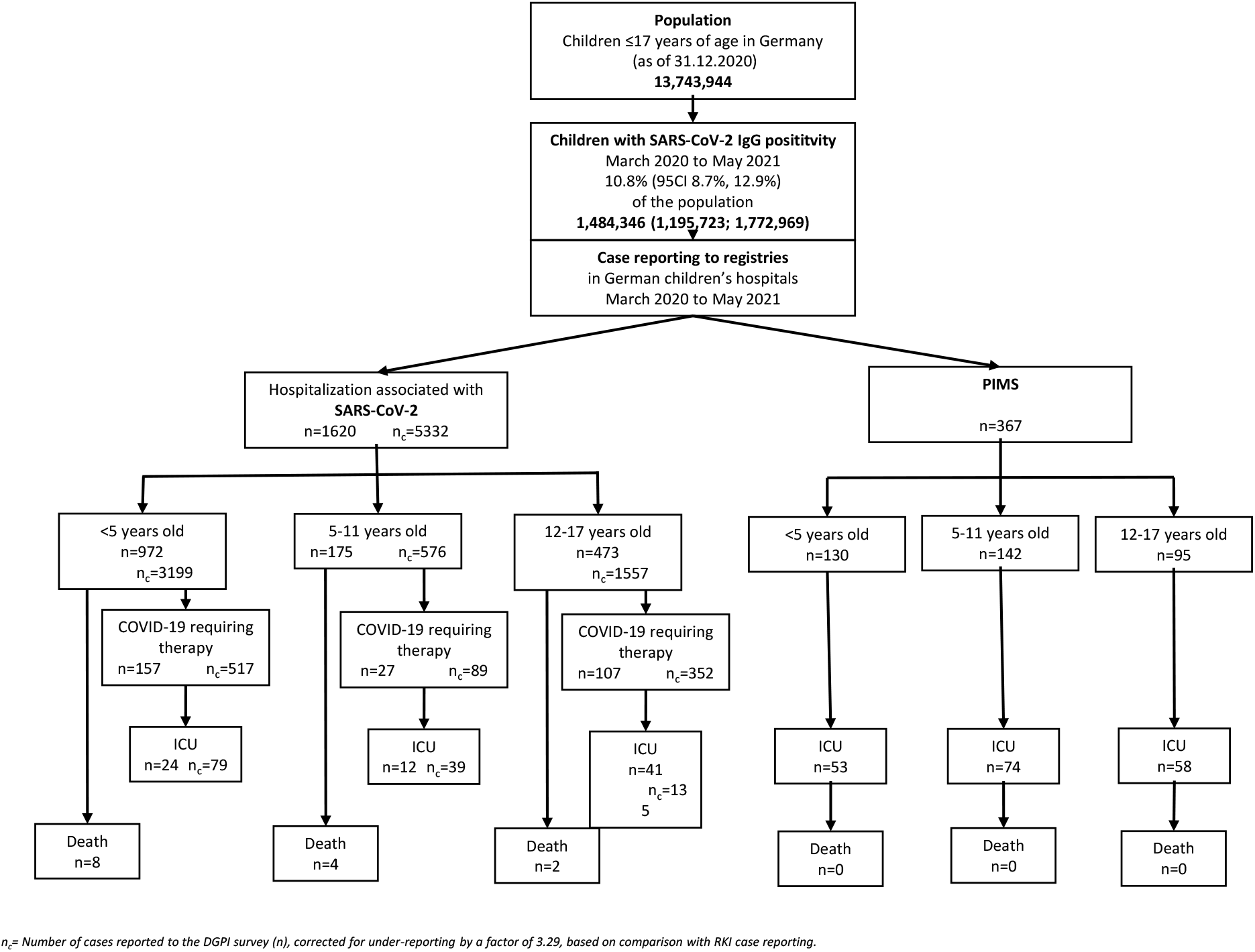

### Risks associated with SARS-CoV-2 infection

Table 1 lists all the risk estimates for hospitalization, therapy-requiring COVID-19, ICU admission due to COVID-19, death due to COVID-19, PIMS-TS and ICU admission due to PIMS-TS. Death due to PIMS-TS is not listed, because none of the reported PIMS-TS cases in Germany died.

**Table 1:**
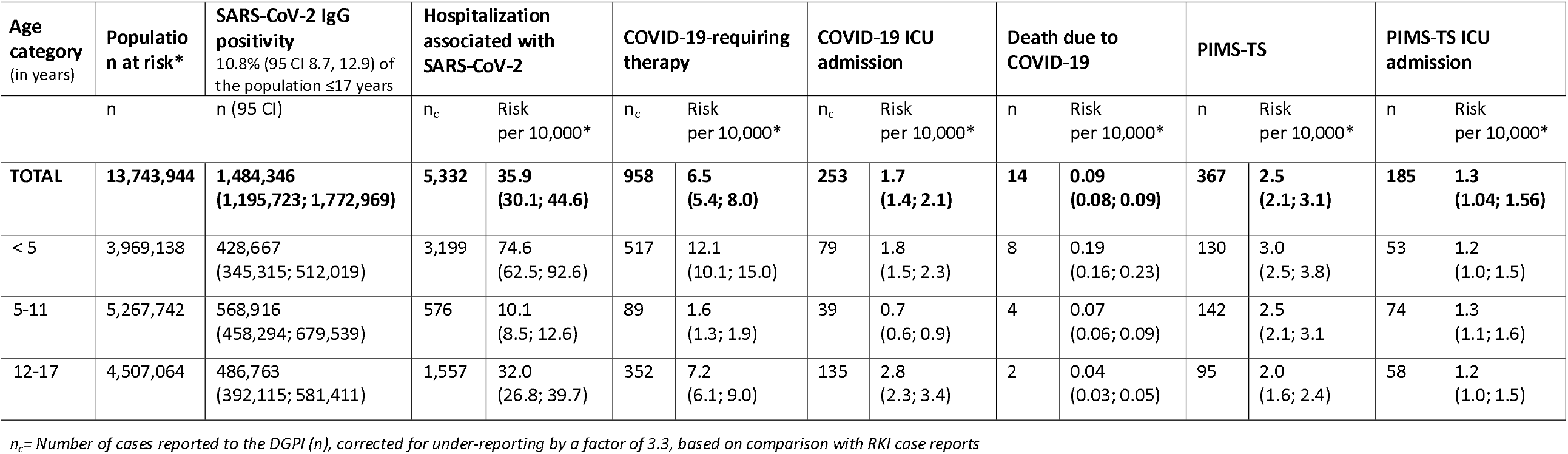
Risk associated with SARS-CoV-2 and PIMS.

| Age category<br>(in years) | Population at risk* | SARS-CoV-2 IgG positivity<br>10.8% (95 CI 8.7, 12.9) of the population ≤17 years | Hospitalization associated with SARS-CoV-2 |  | COVID-19-requiring therapy |  | COVID-19 ICU admission |  | Death due to COVID-19 |  | PIMS-TS |  | PIMS-TS ICU admission |  |
| --- | --- | --- | --- | --- | --- | --- | --- | --- | --- | --- | --- | --- | --- | --- |
|  |  |  | n<sub>c</sub> | Risk per 10,000* | n<sub>c</sub> | Risk per 10,000* | n<sub>c</sub> | Risk per 10,000* | n | Risk per 10,000* | n | Risk per 10,000* | n | Risk per 10,000* |
| **TOTAL** | **13,743,944** | **1,484,346**<br>**(1,195,723; 1,772,969)** | **5,332** | **35.9**<br>**(30.1; 44.6)** | **958** | **6.5**<br>**(5.4; 8.0)** | **253** | **1.7**<br>**(1.4; 2.1)** | **14** | **0.09**<br>**(0.08; 0.09)** | **367** | **2.5**<br>**(2.1; 3.1)** | **185** | **1.3**<br>**(1.04; 1.56)** |
| < 5 | 3,969,138 | 428,667<br>(345,315; 512,019) | 3,199 | 74.6<br>(62.5; 92.6) | 517 | 12.1<br>(10.1; 15.0) | 79 | 1.8<br>(1.5; 2.3) | 8 | 0.19<br>(0.16; 0.23) | 130 | 3.0<br>(2.5; 3.8) | 53 | 1.2<br>(1.0; 1.5) |
| 5-11 | 5,267,742 | 568,916<br>(458,294; 679,539) | 576 | 10.1<br>(8.5; 12.6) | 89 | 1.6<br>(1.3; 1.9) | 39 | 0.7<br>(0.6; 0.9) | 4 | 0.07<br>(0.06; 0.09) | 142 | 2.5<br>(2.1; 3.1) | 74 | 1.3<br>(1.1; 1.6) |
| 12-17 | 4,507,064 | 486,763<br>(392,115; 581,411) | 1,557 | 32.0<br>(26.8; 39.7) | 352 | 7.2<br>(6.1; 9.0) | 135 | 2.8<br>(2.3; 3.4) | 2 | 0.04<br>(0.03; 0.05) | 95 | 2.0<br>(1.6; 2.4) | 58 | 1.2<br>(1.0; 1.5) |

*n<sub>c</sub>*= Number of cases reported to the DGPI (*n*), corrected for under-reporting by a factor of 3.3, based on comparison with RKI case reports

As of May 2021, the cumulative rate for hospitalization associated with SARS-CoV-2 infection was 35.9 per 10,000 children. This rate varied depending upon the seroprevalence, which ranged from 30.1 to 44.6 per 10,000.

When limiting the analysis to include only patients with COVID-19 who required therapeutic interventions, the hospitalization rate decreased 5.5-fold to 6.5 per 10,000 children, and more than 20-fold when the cases counted were limited to those admitted to ICU due to COVID-19 (1.7 per 10,000).

For the outcome measurements of both hospitalization and therapy requiring COVID-19, the rates were highest in the age group <5 years, followed by the age group 12-17 years. By contrast, they were lowest in the age group 5-11 years (both p-values <0.0001). Regarding intensive care treatment, the rate was highest in the 12–17-year age group (p=<0.0001).

Our analysis found a case fatality of 0.09 per 10,000 children through May 2021 in Germany. This was based upon a total number of 14 pediatric fatalities due to COVID-19. The DGPI registry captured almost all of these fatalities, with 13 reported cases as compared to the 14 recorded in the statutory notifications system. In 5/13 (38%) of these cases, the patients had been in palliative care due to an underlying disease prior to their SARS-CoV-2 infection.

Limiting the analysis to children without comorbidities had little impact on the rate of hospitalization. However, with respect to the other outcome measurements – therapy-requiring COVID-19 and ICU admission due to COVID-19 – the estimated risks decreased to 5.1 and 0.8 per 10,000 children, respectively (Table 2). There is a significant association between comorbidity and these outcomes (both p-value <0.0001). Among children without comorbidities, case fatality was 0.03 per 10,000, with no deaths reported in children ≥ 5 years of age.

**Table 1:** Risk associated with SARS-CoV-2 and PIMS for children without comorbidity

| Age category<br>(in years) | Population at risk* | SARS-CoV-2 IgG positivity<br>10.8% (95 CI 8.7, 12.9) of the population ≤17 years | Hospitalization associated with SARS-CoV-2 |  | COVID-19-requiring therapy |  | COVID-19 ICU admission |  | Death due to COVID-19 |  | PIMS-TS |  | PIMS-TS ICU admission |  |
| --- | --- | --- | --- | --- | --- | --- | --- | --- | --- | --- | --- | --- | --- | --- |
|  |  |  | n<sub>c</sub> | Risk per 10,000* | n<sub>c</sub> | Risk per 10,000* | n<sub>c</sub> | Risk per 10,000* | n | Risk per 10,000* | n | Risk per 10,000* | n | Risk per 10,000* |
| **TOTAL** | **9,780,335** | **1,056,276 (850,889; 1,261,663)** | **3,752** | **35.5 (29.7; 44.1)** | **543** | **5.1 (4.3; 6.4)** | **82** | **0.8 (0.7; 1.0)** | **3** | **0.03 (0.02; 0.04)** | **311** | **2.9 (2.5; 3.7)** | **163** | **1.5 (1.3; 1.9)** |
| < 5 | 2,824,480 | 305,044 (245,730; 364,358) | 2,577 | 84.5 (70.7; 104.9) | 362 | 11.9 (9.9; 14.7) | 36 | 1.2 (1.0; 1.5) | 3 | 0.1 (0.08; 0.12) | 117 | 3.8 (3.2; 4.8) | 51 | 1.7 (1.4; 2.1) |
| 5-11 | 3,748,580 | 404,847 (326,126; 483,567) | 342 | 8.5 (7.1; 10.5) | 36 | 0.9 (0.7; 1.1) | 7 | 0.2 (0.1; 0.2) | 0 | - | 121 | 3.0 (2.5; 3.7) | 65 | 1.6 (1.3; 2.0) |
| 12-17 | 3,207,274 | 346,386 (279,033; 413,738) | 833 | 24.1 (20.1; 29.9) | 145 | 4.2 (3.5; 5.2) | 39 | 1.1 (0.9; 1.4) | 0 | - | 73 | 2.1 (1.8; 2.6) | 47 | 1.4 (1.1; 1.7) |

*n<sub>c</sub>* = Number of cases reported to the DGPI (n), corrected for underreporting by a factor of 3.3, based on comparison with RKI case reports
\* Children ≤17 years living in Germany reduced by a proportion of 28.8% (expected maximum number of children with pre-existing conditions, derived from the DGPI data, where 462 out of 1602 children had pre-existing conditions).

For the overall study cohort, the cumulative risk for developing PIMS-TS was 2.5 per 10,000 and 1.3 per 10,000 for ICU admission due to PIMS-TS (Table 1). Significantly fewer cases of PIMS-TS were reported in 12-17-year-old compared to the other age groups (p=0.005), but there were no age differences in ICU admissions due to PIMS-TS (p=0.88). In contrast to the other outcome measurements analyzed, PIMS-TS mainly seems to affect children without comorbidities (p<0.0001). Confining the analysis to children without comorbidities slightly increased the overall risk to 2.9 per 10,000 for PIMS-TS and to 1.5 per 10,00 for ICU admission due to PIMS-TS (Table 2).

## Discussion

By combining the results of a nationwide seroprevalence study [8] together with data from clinical registries and the statutory notification system, we were able to develop reliable estimates of the SARS-CoV-2-associated disease burden, both for the acute infection phase and for PIMS-TS, a hyperinflammatory syndrome that typically usually occurs 4-6 weeks after a mild or asymptomatic SARS-CoV-2 infection.

Overall, the extremely low risk for severe disease course — as indicated by the need for therapeutic intervention, admission to ICU or else death during the acute infection phase — is consistent with data previously reported from other countries [15,16]. Interestingly however, children in the 5-11 age group seem to have a lower risk than children <5 years and adolescents 12-17 years of age.

Overall, the majority of hospitalizations occurred in children and adolescents without comorbidities. Interestingly, however, children with comorbidities account for almost half of all COVID-19 cases requiring therapeutic intervention (415 vs. 543) and two-thirds of cases requiring ICU admission (171 vs. 82), even though they make up only a small portion of the pediatric population. Primarily healthy children ages 5-11 have the lowest risk with a 0.2/10,000 rate of ICU admission. Due to an absence of cases, their case fatality rate cannot be calculated.

Previously, the only existing estimates regarding the true incidence of PIMS-TS after a SARS-CoV-2 infection were based on rough estimates of the number of infections. With our data, however, we were able to calculate an overall risk of 1/4,000 cases. With approximately 50% of these cases requiring ICU admission, PIMS-TS plays a relevant role in the overall SARS-CoV-2-associated disease burden in the pediatric age group. It leads to approximately one quarter of all therapy-requiring hospitalizations and almost 40% of all ICU admission. In children without comorbidities, the role of PIMS-TS is even more pronounced and accounts for 38% of all therapy-requiring hospitalizations and 65% of ICU admissions.

Fortunately, there are effective treatment options for PIMS-TS. Only small numbers of patients have suffered from sequalae and follow-up data emerging from the UK and the US has been reassuring [17–19]. In addition, both national [11] and international [20] PIMS-TS registries seem to be detecting a lower rate of PIMS-TS cases, as the delta-variant has predominated. While this data is still preliminary and needs to be validated, these preliminary observations are important to consider in future analyses, especially given the role that PIMS-TS plays in the overall SARS-CoV-2 associated disease burden among children and adolescents.

### Strengths and limitations

The strength of our analysis lies in its provision of reliable estimates for evaluating the risk of severe manifestations related to SARS-CoV-2 infections in children.

Our analysis has several limitations, mainly due to uncertainties in the raw data in the three different sources, which are mostly estimates.

The population at risk: We do not know the exact number of SARS-CoV-2 infections in children in Germany. We have estimated the number based upon recently published results of a nationwide, prospective survey of IgG antibodies in pediatric patients in German pediatric hospitals. Limitations include potential selection bias, ELISA cut-off values, precision and the suitability of IgG detected by ELISA to depict SARS-CoV-2 infections in children. Nevertheless, the data are comparable to the standards of other seroprevalence estimates worldwide. On this basis, our data provide the best available estimates for Germany.

The number of cases: Reporting to the DGPI register is voluntary, while reporting laboratory-confirmed SARS-CoV-2 infections to the statutory notification system is compulsory. Thus, the latter may be assumed to be more comprehensive. Because case definitions in the DGPI register and the statutory notification system were identical, this provides a reasonable basis for correcting the underreporting of the DGPI cases based upon the statutory notification system. Limiting the analysis to children up to the age of 14 (rather than 17) is a minor limitation.

As in all voluntary reporting systems, reporting may be biased towards more severe cases. This presumption is supported by the fact that only 30% of all hospitalizations were captured, while more than 90% of deaths were reported. By correcting therapy-requiring COVID-19 and ICU admissions based upon the factor calculated for overall hospitalizations, we likely overestimated the true SARS-CoV-2-associated burden to some degree. The inclusion of patients with concurrent viral and bacterial infections also may have added to this overestimation. However, the overall numbers of these patients were low. On the other hand, we suspect there may have been a degree of underreporting in the PIMS-TS registry. This underreporting could lead to an underestimation of risks associated with PIMS-TS. No correction was feasible for PIMS-TS. However, because PIMS-TS is a newly discovered, severe condition requiring hospitalization, it seems reasonable to assume that reporting will become more comprehensive over time. As a point of comparison, in a different but similar, active, non-compulsory, pediatric, hospital-based surveillance system in Germany, meningitis was more likely to be comprehensively reported than a positive blood culture result [21].

Our identification of relevant cases based upon a case definition that specifically requires COVID-19-related treatment critically influences the numbers present in our analysis. However, reporting physicians are the ones in the best position to assess their patients and identify appropriate therapeutic interventions for SARS-CoV-2 infections. These patients may have been admitted to hospital for other medical conditions. In these instances, they should not be counted towards the SARS-CoV-2-associated disease burden in their age group.

The applied estimate of the proportion of children without comorbidities was chosen in order to avoid underestimation. According to a report by Schröder et al., which was based upon German health insurance data, only approximately 3% of all children in Germany have one or more relevant comorbidity and — accordingly — an increased risk for a severe COVID-19 course [22]. Finally, the precision of our estimates is difficult to measure. We used the 95% confidence limits of the estimate of the population at risk in order to capture the uncertainties. We disregarded the uncertainties in the number of cases, because the number was corrected according to the presumed true number of cases provided by the statutory notification system.

### Conclusions

Our analysis of the data shows that the SARS-CoV-2-associated burden of severe disease or death in children and adolescents is low. This is particularly true for COVID-19 related symptoms in 5-11-year-old children without comorbidities. PIMS-TS appears to play a crucial role in overall SARS-CoV-2 associated disease burden and exceeds the risks of the acute severe COVID-19 children and adolescents without comorbidities.

## Data Availability

We share data if reasonable requests are received. Requests should be directed to the corresponding author at

## Acknowledgments

We thank each and every staff member in the participating hospitals for reporting their cases to the DGPI registry.

## Notes

### Competing Interest Statement

The authors have declared no competing interest.

### Funding Statement

This study did not receive any funding

### Author Declarations

The DGPI registry was approved by the Ethics Committee of the Technische Universitaet (TU) Dresden (BO-EK-110032020) and was assigned clinical trial number DRKS00021506.

